# Retention of Neutralizing response against SARS-CoV-2 Omicron variant in Sputnik V vaccinated individuals

**DOI:** 10.1101/2022.01.15.22269335

**Authors:** D Lapa, DM Grousova, G Matusali, S Meschi, F Colavita, A Bettini, G Gramigna, M Francalancia, AR Garbuglia, E Girardi, V Puro, A Antinori, AV Kovyrshina, IV Dolzhikova, DV Shcheblyakov, AI Tukhvatulin, OV Zubkova, DY Logunov, BS Naroditsky, F Vaia, AL Gintsburg

## Abstract

The new variant Omicron (B.1.1.529) of SARS-CoV-2, first identified in November 2021, is rapidly spreading all around the world. The Omicron becomes the dominant variant of SARS-CoV-2. There are many ongoing studies evaluating the effectiveness of existing vaccines. Studies on neutralizing activity of vaccinated sera against Omicron variant are currently being carried out in many laboratories.

In this study, we have shown the neutralizing activity of sera against SARS-CoV-2 Omicron (B.1.1.529) variant compared to the reference Wuhan D614G (B.1) variant in individuals vaccinated with 2 doses of Sputnik V or BNT162b2 in different time points up to 6 months after vaccination. We performed analysis on sample pools with comparable NtAb to Wuhan D614G variant. The decrease in neutralizing antibody (NtAb) to the Omicron variant was 8.1 folds for group of Sputnik V-vaccinated and 21.4 folds for group of BNT162b2-vaccinated. Analysis showed that 74.2% of Sputnik V- and 56.9% of BNT162b2-vaccinated sera had detectable NtAb to SARS-CoV-2 Omicron variant.

The decrease in NtAb to SARS-CoV-2 Omicron variant compared to Wuhan variant has been shown for many COVID-19 vaccines in use, with some showing no neutralization at all. Today the necessity of third booster vaccination is obvious. And the most effective approach, already shown in several studies, is the use of heterologous booster vaccination pioneered in COVID-19 vaccines by Sputnik V.

## Introduction

As of Jan 12, 2022, the Omicron variant has been confirmed in at least 150 countries around the world [1]. From Jan 03, 2022 to Jan 09, 2022 Omicron variant accounts for roughly 62.5 percent of all SARS-CoV-2 sequences available on GISAID database [2].

Today, the effectiveness of existing vaccines against the new Omicron variant is the question of high priority. Around the world, studies are being carried out to analyze the neutralization activity of the sera of vaccinated people against the Omicron variant.

It was shown, that sera from vaccinated individuals with 2 doses BNT162b2 or mRNA-1273 has 22-30-fold decrease in neutralizing activity against the Omicron variant compared with wild type virus [3-4]. A similar decrease in neutralizing activity was shown in other study comparing two vaccines: ChAdOx1-nCoV-19 and BNT162b2 [4-5]. A study was conducted in China comparing the effectiveness of the BNT162b2 and Coronavac vaccines. According to the results of the study, the complete no neutralizing antibodies against the Omicron variant was shown in individuals vaccinated with Coronavac [6].

However, many studies have shown that administration of 3 doses of vaccine or a combination of disease and vaccination significantly increases the titer of virus-neutralizing antibodies to Omicron variant [7-8]. Analysis of sera of individuals only vaccinated with BNT162b2, mRNA-1273 or Ad26.COV2.S shown the decrease in neutralizing activity against the Omicron variant from 23 to 122 folds. In case of combination of infection and vaccination, the decrease in titers was from 4 to 13 folds [9].

In this study, we have shown the neutralizing activity of sera against Omicron variant in individuals vaccinated with 2 doses of Sputnik V or BNT162b2 in different time points after vaccination and in Sputnik V-vaccinated individuals with a history of COVID-19.

## Results

We carried out a study of the neutralizing activity of the blood sera of the Sputnik V and BNT162b2 vaccinated individuals against Omicron and Wuhan D614G variant of SARS-CoV-2. In vaccinated group we included individuals who were vaccinated with two doses of Sputnik V (n = 31) or two doses of BNT162b2 (51 samples from 17 individuals) and with no history of COVID-19 (absence of antibodies to SARS-CoV-2 N-protein, no report of SARS-CoV-2 positive molecular test). The descriptions of the groups are presented in tables S1 and S2. The median and IQR of time after the second dose of the vaccine was 91 days (56-122) for Sputnik V and 90 days (14-180) for BNT162b2 (p = 0.8117, Mann-Whitney test). Analysis of RBD-specific antibodies showed that antigen-specific IgG was detected in all samples (Figure 1A), the level of IgG between the Sputnik V and BNT162b2 groups was comparable (p = 0.0801, Mann-Whitney test). Notably, IgG dynamics was different in groups: Sputnik V sera had stable IgG level in time while samples from BNT162b2-vaccinated showed peak of IgG response on 2 weeks with significant decrease at 3 and 6 months timepoints (Figure S1). Analysis of neutralizing antibodies (NtAb) to the Wuhan D614G variant also did not show statistically significant differences between the groups (Figure 1B, p = 0.2658, Mann-Whitney test). So we performed analysis on sample pools with comparable RBD-specific IgG and NtAb to Wuhan D614G variant. The decrease in the NtAb level to the Omicron variant in comparison to the B.1 variant in the Sputnik V vaccinated sera was 8.1 folds (GMT 58.5 vs 7.2), in the sera of individuals vaccinated with BNT162b2 - 21.4 folds (GMT 72.7 vs 3.4). In general, Omicron-specific NtAbs were detected in the blood sera of 74.2% of the Sputnik V-vaccinated, and 56.9% of the BNT162b2-vaccinated (Figure 1C, 1D).

**Figure 1.**
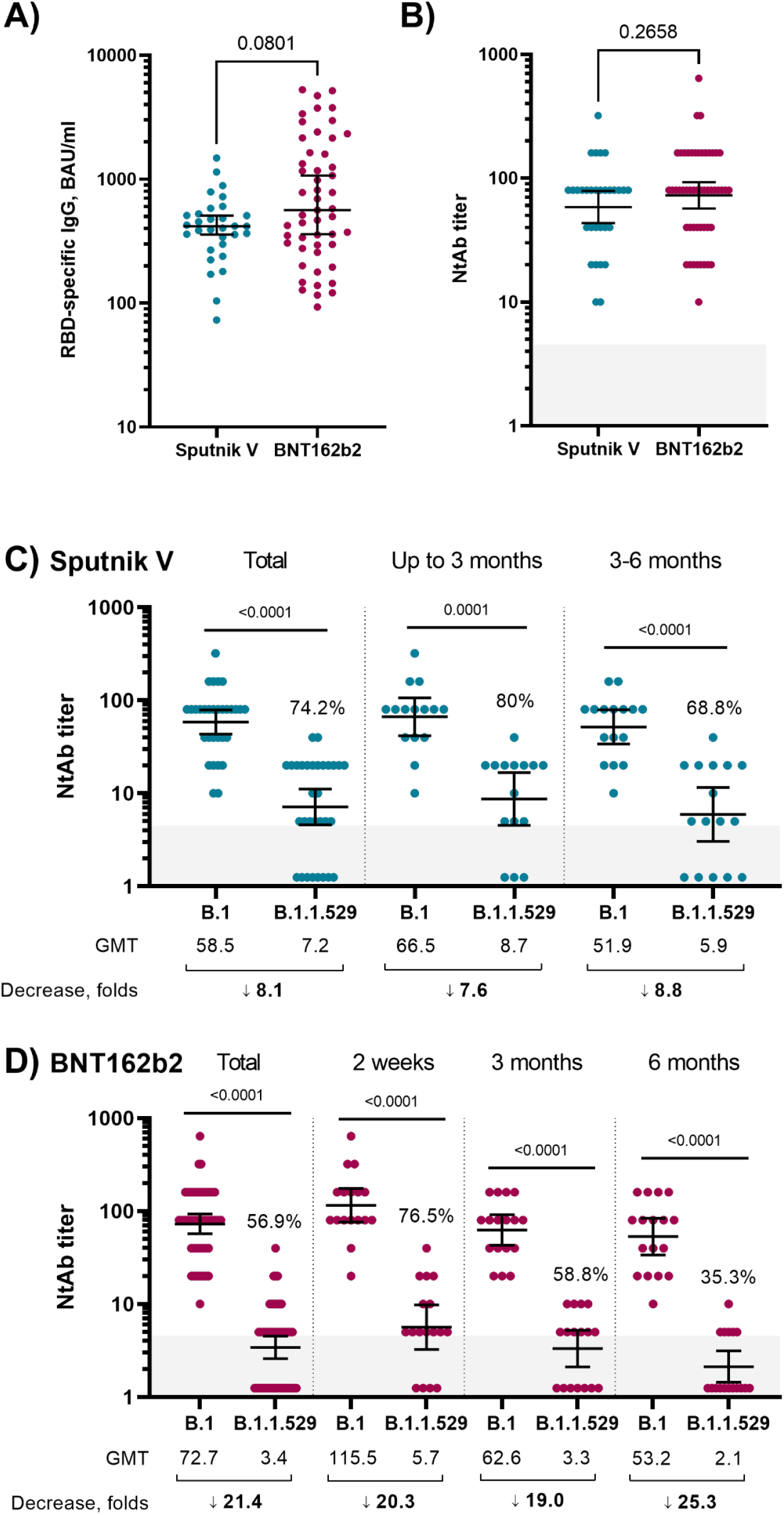
A –RBD-specific IgG titers in vaccinated individuals, median and 95% CI,; B – NtAb titers against SARS-CoV-2 B.1 in vaccinated individuals, geometric mean titer and 95% CI, C, D - NtAb titers against SARS-CoV-2 B.1 (Wuhan D614G) and B.1.1.529 (Omicron) variants in Sputnik V (C) and BNT162b2 (D) vaccinated individuals, geometric mean titer and 95% CI, % - part of sera with detectable NtAb. P-value was determined by Wilcoxon test. Grey box – limit of detection. Values below the limit of detection were assigned a value of NtAb 1.25.

Next, we stratified the samples based on the time period after receiving the second dose of vaccines (Figure 1C, 1D). In the sera of individuals vaccinated with second dose less than 3 months ago with Sputnik V (n = 15), the decrease in NtAb to the Omicron variant was 7.6-fold, NtAbs to the Omicron variant were detected in 80% of the samples. Samples from individuals who received the second dose of Sputnik V 3-6 months ago (n = 16), the NtAb titers showed 8.8-fold decrease (NtAbs were detected in 68.8% of samples) (Figure 1C). Analysis of samples from individuals vaccinated with BNT162b2 (Figure 1D) showed that 2 weeks after the 2nd dose, the was 20.3-fold decrease in NtAbs to the Omicron variant (NtAbs to Omicron variant were detected in 76.5% of samples). In subgroups of 3 and 6 months after the second dose the decrease in NtAb titer was 19- and 25.3-fold, respectively, while NtAbs to Omicron were detected in 58.8% and 35.3% samples, respectively (Figure 1D).

We also analyzed the NtAb level in the blood sera of individuals who were vaccinated with Sputnik V and underwent COVID-19 (before or after vaccination) in an asymptomatic form (there is no history of COVID-19, but there are antibodies specific to the N protein) and symptomatic form (medical record of mild/moderate COVID-19) and in COVID-19 convalescent. In the sera of people vaccinated with Sputnik V and undergoing asymptomatic COVID-19 (n = 8) and symptomatic COVID-19 (n = 12), the decrease in NtAb titers against the Omicron variant was 6.7-fold (GMT 160 vs 23.8, NtAbs to Omicron were detected in 87.5 % of samples) and 5.0-fold (GMT 119.9 vs 23.8, NtAbs to Omicron detected in 100% of samples), respectively. Analysis of convalescent samples (n=23) showed comparable decrease to Omicron variant (6.0 folds and 60.9% responders) (Figure 2).

**Figure 2.**
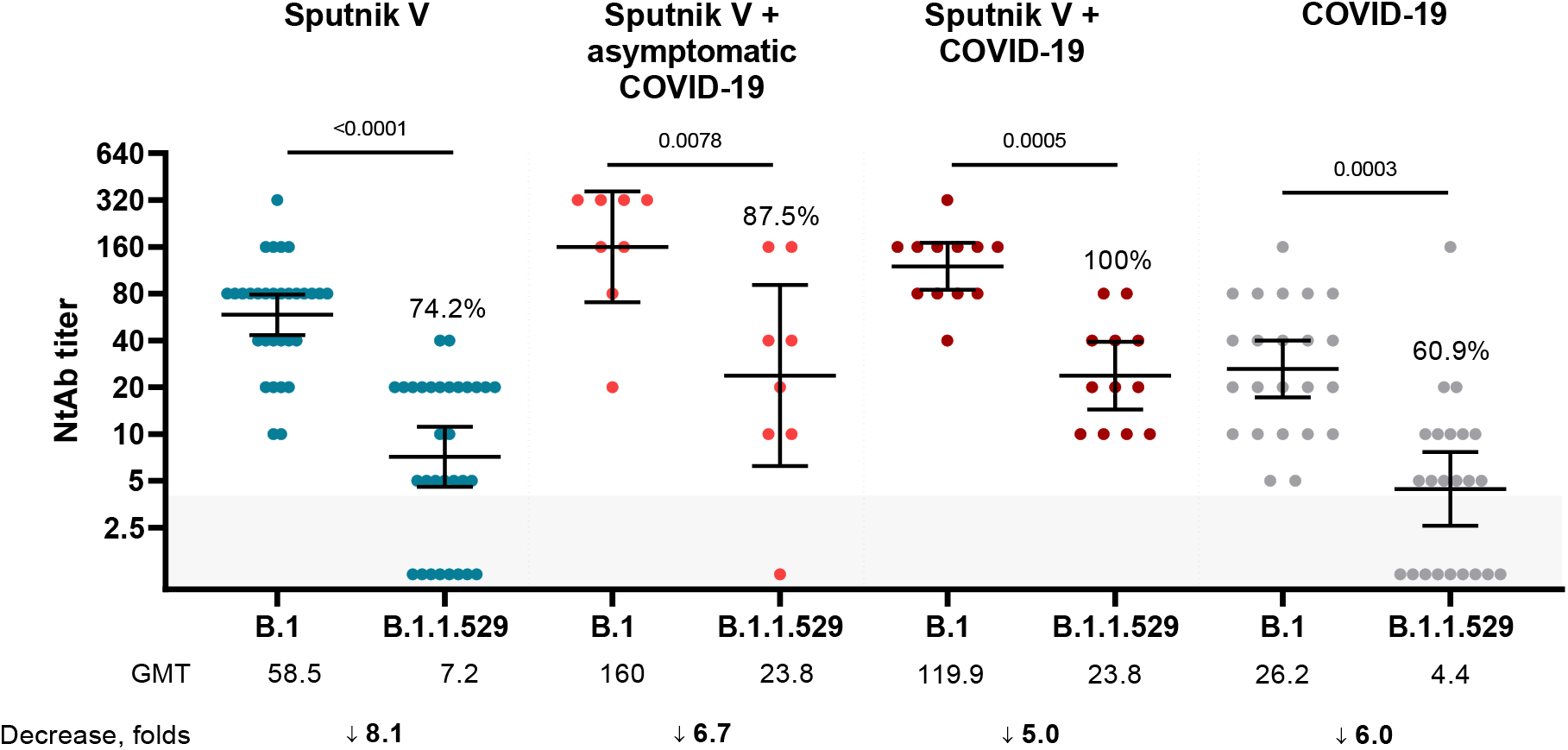
Titers of neutralizing antibodies in samples of individuals vaccinated with Sputnik V; vaccinated with Sputnik V after/before asymptomatic COVID-19; vaccinated with Sputnik V after/before mild COVID-19 and COVID-19 convalescents. The figure shows individual data for the studied samples, geometric mean and 95% CI. The figure shows the p-value (Wilcoxon test), % of individuals with detectable NtAb to the SARS-CoV-2, geometric mean and the level of NtAb decrease. Grey box – limit of detection. Values below the limit of detection were assigned a value of NtAb 1.25.

## Discussion

In this study we investigated neutralizing activity of sera of two-dose Sputnik V-vaccinated individuals. For this purpose, we performed live-virus neutralization assay with SARS-CoV-2 Wuhan (B.1) and Omicron (B.1.1.529) variants. As a control to our study we also assessed the ability of serum samples from BNT162b2 (two doses) vaccinated health care workers to neutralize the Omicron variant. Initially both pools of vaccinated sera (total pool of different timepoints) had comparable RBD-specific IgG and NtAb to Wuhan D614G SARS-CoV-2 variant. According to the results of the study, fold decrease of virus-neutralizing antibodies to Omicron vs Wuhan variant in the group of Sputnik V-vaccinees was 8.1, while, in line with recent publications [3-6], in BNT162b2-vaccinated group we observed a 21.4 fold reduction.

We divided each group of vaccinated sera into 7 parts according to RBD-specific IgG (BAU/ml) quartiles (0-25%, >25-50%, >50-75%, >75-100%, 0-50%, >25-75% and > 50-100%) and counted decrease of NtAb and percent of sera with detectable NtAb (Table S4). We have shown similar levels of neutralization drop among different groups, divided on the basis of quartiles (Table S4).

We also analyzed neutralizing activity of sera of Sputnik V-vaccinated individuals with a history of mild or asymptomatic COVID-19. The decrease of NtAb titers to Omicron in the groups of Sputnik V-vaccinated COVID-19 convalescents both mild and asymptomatic was 5.0-fold and 6.7-fold respectively. In both groups fold decrease of NtAb titers was lower than in vaccinated groups and GMT against Omicron variant was 23.8, while GMT in vaccinated groups didn’t exceed 7.0. Interestingly the decrease of NtAb in Sputnik V and Sputnik V + COVID-19 samples was similar to what was shown in samples from unvaccinated-COVID-19 convalescent individuals where a 6.0-fold decrease in the NtAb titer against Omicron was observed.

Sputnik V shows the promising results: after 2 doses, there is an 8.1-fold decrease in the neutralizing activity of sera against the Omicron variant, which is significantly less than the decrease for other vaccines after 2 doses. The advantages of Sputnik V are the use of native S glycoprotein (without proline-stabilization and other modifications that may move an immune response predominantly to the actively mutating receptor-binding domain of S glycoprotein – the Omicron variant has 16 mutations in RBD) and the use of a heterologous prime-boost vaccination regimen. This approach allows to form a robust and long-lasting immune response after only 2 doses: this study showed that NtAb to the Omicron variant were detected in 68.8% of Sputnik V sera 3-6 months after vaccination, previous study showed that 55.6% of vaccinated sera had NtAb to Omicron variant 6-12 months after vaccination [9].

This study shows data on neutralizing activity of Sputnik V-vaccinated and BNT162b2-vaccinated sera to B.1.1.529 (Omicron) SARS-CoV-2 variant. This study has several limitations: 1) the relatively small number of tested samples in time groups, which may not reflect the dynamics of neutralizing antibodies; 2) absence of data about age-dependent and comorbidity-dependent NtAb response; 3) some samples with moderate virus-neutralizing antibody titers on Wuhan D614G variant are followed by a number of results on Omicron variant being under the limit of detection and hence limited accuracy of evaluation of virus-neutralizing antibody titers decrease; 4) heterogeneity of vaccinated convalescents group.

The results have shown that two-dose vaccination regimen allows the formation of neutralizing antibodies that can neutralize the Omicron variant, and we suppose that it may be sufficient in protection against COVID-19-associated hospitalizations and deaths. However, it is unclear whether this level of antibodies will be enough to effectively neutralize the Omicron variant to protect against infection. This study together with previous studies gives additional evidence for the need of booster immunization [9-13]: Sputnik Light booster after Sputnik V vaccination induces robust neutralizing antibody response to Omicron variant [9]. Immune response to SARS-CoV-2 Omicron variant after booster immunization is currently being actively investigated [9-13]. Vaccine research shows that vaccinated individuals have a significantly milder course of Omicron variant infection. And the strategy of using booster doses allows to reduce not only severity, but also the frequency of COVID-19 cases caused by the Omicron variant [10]. In the scope of vaccination strategies against new SARS-CoV-2 variants, emerging evidence indicates that heterologous vaccination could improve the immune response as compared to homologous vaccination [13-15]: Ad26.COV2.S enhances the immunogenicity of BNT162b2, especially to the Omicron variant [13].

## Materials and methods

### Ethics statement

The study was conducted according to the guidelines of the Declaration of Helsinki, and approved by the INMI Ethical Committee (issue n. 297/2021) and Gamaleya NRCEM Local Ethics Committee (Protocol No. 17 of December 03, 2021).

### Samples for analysis

Peripheral blood was collected from Sputnik V vaccinated, BNT162b2 vaccinated, and from COVID-19 convalescent individuals (Tables S1, S2 and S3). Part of COVID-19 convalescent samples were plasma samples. Serum/plasma samples were centrifuged at 1000g for 10 minutes, aliquoted and stored at -80°C until use.

### RBD- and N-specific IgG Evaluation

Two commercial chemiluminescence microparticle antibody assays (ARCHITECT, Abbott Laboratories, Wiesbaden, Germany) were used, according to manufacturer’s protocols: the anti-nucleoprotein IgG and the SARS-CoV-2 IgG II kit, which detected antibodies against the RBD of SARS-CoV-2. Index values≥ 1.4 and values≥ 7.1 BAU/mL are considered positive for anti-N IgG and anti-RBD IgG respectively.

### SARS-CoV-2 variants

As a Wuhan D614G reference strain we used isolate B.1 (SARS-CoV-2/Human/ITA/PAVIA10734/2020, EVAG Ref-SKU 008V-04005, GISAID accession ID EPI_ISL_568579). The Omicron variant of SARS-CoV-2 was isolated from a nasopharyngeal swab of a patient who arrived from South Africa to Italy in December 2021(hCoV-19/Italy/LAZ-INMI-2890/2021, GISAID accession ID EPI_ISL_7716384).

### Cell culture, virus isolation and propagation

Viral isolation was performed on Vero E6/TMPRSS2 (kindly provided by Dr. Oeda S., National Institute of Infectious Diseases, Tokyo, Japan). Initial passage, propagation and titration were performed on Vero E6 cells (ATCC CRL-1586). Cells were maintained in Minimal Essential Medium (MEM), containing 10% heat-inactivated fetal bovine serum (Corning), L-glutamine (Corning) and penicillin/streptomycin solution (Corning).

Virus titre was determined by limiting dilution assay and residual infectivity was expressed as 50% Tissue Culture Infective Dose (TCID50/ml) calculated according to the Reed and Muench method. All work with infectious SARSCoV-2 virus was performed under biosafety level 3 (BSL-3) conditions at INMI.

### Neutralization assay

Neutralizing antibodies against SARS-CoV-2 variants in sera/plasma samples were analyzed by microneutralization test. The assay was performed according to described earlier [16], using B.1 and VOC B.1.1.529 as challenging viruses. Briefly, serum/plasma samples after heat inactivation (+56 °C for 30 minutes) were serially diluted in MEM supplemented with 2% HI-FBS with starting sample dilution at 1:5 with two-fold dilution and mixed with 100 TCID50 SARS-CoV-2 at 1:1 ratio (50 μl serum dilution and 50 μl virus suspension) and incubated at 37 °C for 30 minutes. After that, serum-virus complexes were transferred to Vero E6 cells in 96-well plates and incubated for 48 h for B.1. and for 72 h for B.1.1.529 (Omicron). The 90% cytopathic effect (CPE) was assessed visually, if even a slight damage to the monolayer (1-3 "plaques") was observed in the well. Wells with more damage to the monolayer (4 and more "plaques”) was considered as a well with a manifestation of CPE. Neutralization titer was defined as the highest serum dilution with 90% CPE or without CPE in two replicable wells. To standardize the inter-assay procedures, positive control samples showing high and low neutralizing activity were included in each session.

### Statistical analysis

Statistical analysis was performed in GraphPad Prism version 9.2.0 (GraphPad Software Inc., San Diego, CA, USA). For comparison of paired data, Wilcoxon test was used, for comparison of unpaired data – Mann-Whitney test.

## Data Availability

All data produced in the present work are contained in the manuscript

## Supplementary

**Table S1.** Summary table of participants vaccinated with Sputnik V

|  | Sputnik V<br>(up to 6 months) | Sputnik V +<br>asymptomatic* COVID-19 | Sputnik V + mild<br>COVID-19 |
| --- | --- | --- | --- |
| Number of individuals | 31 | 8 | 12 |
| Age, years** | 33 (25-47) | 38.5 (27.25-44.75) | 36 (30.25-42.5) |
| Male sex | 16 (51.6%) | 6 (75%) | 11 (91.7%) |
| RBD-specific IgG, BAU/ml ** | 416.6 (298.8-525.9) | 878.1 (390.1-2772) | 767.2 (516.5-1459) |

\* No COVID-19 in anamnesis, SARS-CoV-2 N-protein positive IgG.
\*\* values are median (interquartile range).

**Table S2.** Summary table of participants vaccinated with BNT162b2 (2w, 3m, 6m)

|  | BNT162b2 |
| --- | --- |
| Number of individuals | 17 |
| Age, years* | 37 (31-48) |
| Male sex | 9 (53%) |
| RBD-specific IgG, BAU/ml * | 562.7 (294.9-1640) |

\* values are median (interquartile range).

**Table S3.**
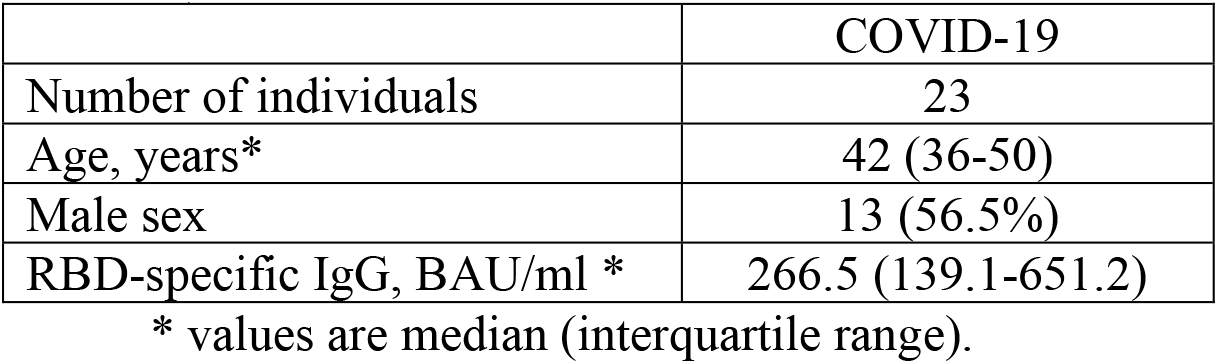
Summary table of COVID-19 convalescent participants (samples collected at 1 to 3 months from symptom onset in march to December 2020, Italy and in January to March 2021 in Russia)

|  | COVID-19 |
| --- | --- |
| Number of individuals | 23 |
| Age, years* | 42 (36-50) |
| Male sex | 13 (56.5%) |
| RBD-specific IgG, BAU/ml * | 266.5 (139.1-651.2) |

\* values are median (interquartile range).

**Table S4.** Decrease of NtAb to Omicron variant vs B.1 variant in sera of vaccinated according to basic level of RBD-specific IgG

| Group | Sputnik V |  | BNT162b2 |  |
| --- | --- | --- | --- | --- |
|  | NtAb decrease, folds | % of sera with detectable NtAb | NtAb decrease, folds | % of sera with detectable NtAb |
| Total | 8.1 | 74.2 | 21.4 | 56.9 |
| 0-25% | 10.4 | 25 | 24.5 | 15.4 |
| >25-50% | 6.2 | 87.5 | 19.8 | 46.2 |
| >50-75% | 11.3 | 87.5 | 16.9 | 76.9 |
| >75-100% | 5.9 | 100 | 25.4 | 83.3 |
| 0-50% | 8.0 | 56.3 | 22.0 | 34.6 |
| >50-100% | 8.4 | 93.3 | 20.5 | 80 |
| >25-75% | 8.4 | 87.5 | 18.3 | 65.4 |

**Figure S1.**
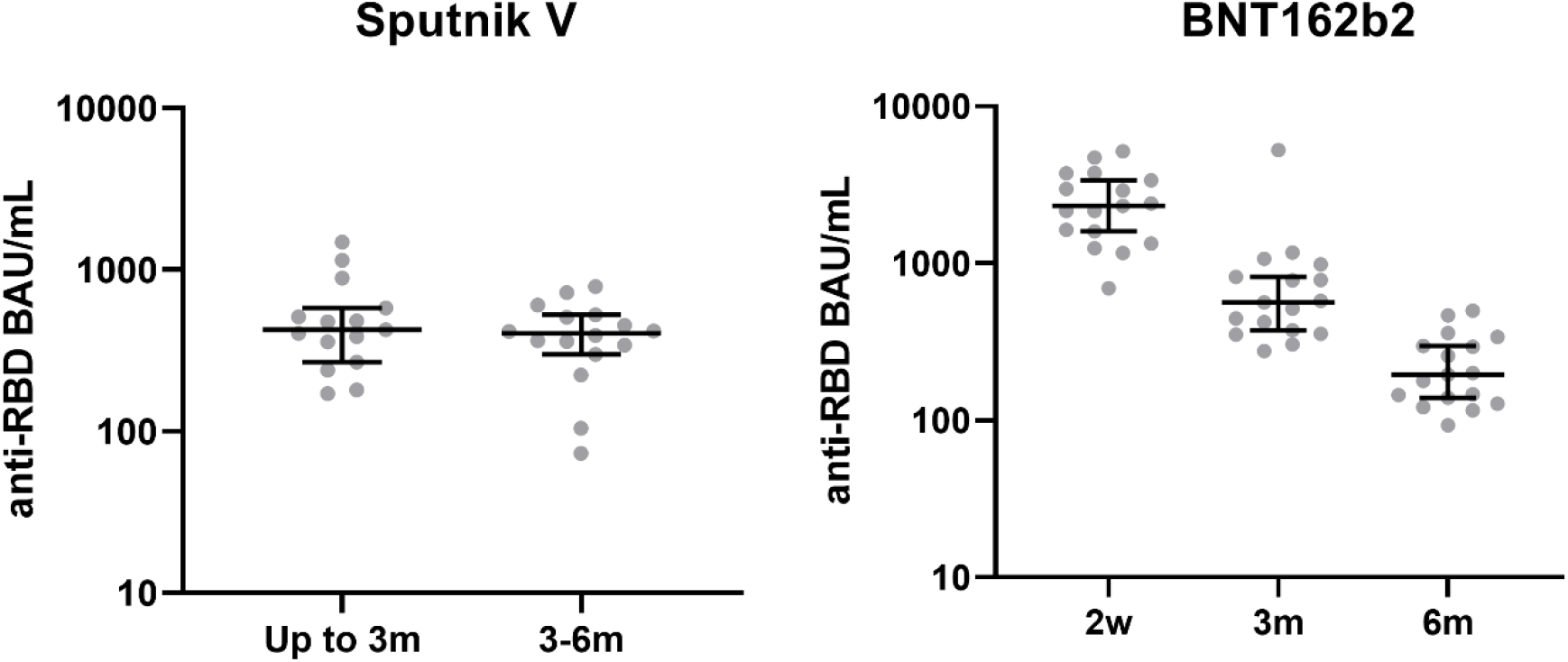
RBD-specific IgG titers in vaccinated individuals, median and 95% CI. W – weeks, m – months.

## References

1. NewsNodes. Omicron Variant Tracker. URL: https://newsnodes.com/omicron_tracker/12.01.2022

2. GISAID. hCov19 Variants. URL: https://www.gisaid.org/hcov19-variants/12.01.2022

3. Edara VV, Manning KE, Ellis M, et al. mRNA-1273 and BNT162b2 mRNA vaccines have reduced neutralizing activity against the SARS-CoV-2 Omicron variant. Preprint. bioRxiv. 2021;2021.12.20.473557. Published 2021 Dec 22. doi:10.1101/2021.12.20.473557

4. Alexander Muik, Bonny Gaby Lui, Ann-Kathrin Wallisch, et al. Neutralization of SARS-CoV-2 Omicron pseudovirus by BNT162b2 vaccine-elicited human sera. medRxiv 2021.12.22.21268103; doi: https://doi.org/10.1101/2021.12.22.21268103

5. Dejnirattisai W, Shaw RH, Supasa P, et al. Reduced neutralisation of SARS-CoV-2 omicron B.1.1.529 variant by post-immunisation serum [published online ahead of print, 2021 Dec 20]. Lancet. 2021;S0140-6736(21)02844-0. doi:10.1016/S0140-6736(21)02844-0]

6. Lu L, Mok BW, Chen LL, et al. Neutralization of SARS-CoV-2 Omicron variant by sera from BNT162b2 or Coronavac vaccine recipients [published online ahead of print, 2021 Dec 16]. Clin Infect Dis. 2021;ciab1041. doi:10.1093/cid/ciab1041

7. Schmidt F, Muecksch F, Weisblum Y, et al. Plasma neutralization properties of the SARS-CoV-2 Omicron variant. Preprint. medRxiv. 2021;2021.12.12.21267646. Published 2021 Dec 13. doi:10.1101/2021.12.12.21267646

8. Doria-Rose NA, Shen X, Schmidt SD, et al. Booster of mRNA-1273 Vaccine Reduces SARS-CoV-2 Omicron Escape from Neutralizing Antibodies. Preprint. medRxiv. 2021;2021.12.15.21267805. Published 2021 Dec 15. doi:10.1101/2021.12.15.21267805

9. IV Dolzhikova, AA Iliukhina, AV Kovyrshina, et al. Sputnik Light booster after Sputnik V vaccination induces robust neutralizing antibody response to B.1.1.529 (Omicron) SARS-CoV-2 variant Preprint. medRxiv. 2021; Published 2021 Dec 21. doi: 10.1101/2021.12.17.21267976

10. UK Health Security Agency. SARS-CoV-2 variants of concern and variants under investigation in England. Technical briefing: Update on hospitalisation and vaccine effectiveness for Omicron VOC-21NOV-01 (B.1.1.529). December 31, 2021.

11. Garcia-Beltran WF, St Denis KJ, Hoelzemer A, et al. mRNA-based COVID-19 vaccine boosters induce neutralizing immunity against SARS-CoV-2 Omicron variant. Cell. 2022 Jan 6:S0092-8674(21)01496-3. doi: 10.1016/j.cell.2021.12.033. Epub ahead of print. PMID: 34995482; PMCID: PMC8733787.

12. Ai J, Zhang H, Zhang Y, et al. Omicron variant showed lower neutralizing sensitivity than other SARS-CoV-2 variants to immune sera elicited by vaccines after boost. Emerg Microbes Infect. 2021 Dec 22:1–24. doi: 10.1080/22221751.2021.2022440. Epub ahead of print. PMID: 34935594

13. C. Sabrina Tan, Ai-ris Y. Collier, Jinyan Liu, et al. Homologous and Heterologous Vaccine Boost Strategies for Humoral and Cellular Immunologic Coverage of the SARS-CoV-2 Omicron Variant. medRxiv 2021.12.02.21267198; doi: https://doi.org/10.1101/2021.12.02.21267198

14. European Medicines Agency. Heterologous primary and booster COVID-19 vaccination. URL: https://www.ema.europa.eu/en/documents/report/heterologous-primary-booster-covid-19-vaccination-evidence-based-regulatory-considerations_en.pdf

15. Sablerolles RSG, Goorhuis A, GeurtsvanKessel CH, et al. Heterologous Ad26.COV2.S Prime and mRNA-Based Boost COVID-19 Vaccination Regimens: The SWITCH Trial Protocol. Front Immunol. 2021 Sep 24;12:753319. doi: 10.3389/fimmu.2021.753319. PMID: 34691071; PMCID: PMC8529966.

16. Meschi, S.; Colavita, F.; Bordi, L. Performance evaluation of abbott architect SARS-CoV-2 IgG immunoassay in comparison with indirect immunofluorescence and virus microneutralization test. J. Clin. Virol. 2020, 129, 104539

